# Frailty is a predictor of medication-related harm requiring healthcare utilisation: a multicentre prospective cohort study

**DOI:** 10.1101/2021.05.17.21257344

**Authors:** Jennifer M. Stevenson, Nikesh Parekh, Kia-Chong Chua, J. Graham Davies, Rebekah Schiff, Chakravarthi Rajkumar, Khalid Ali

**Author notes:** **Corresponding author:** Professor C Rajkumar Address : Brighton and Sussex Medical School, Audery Emerton Building, RSCH, Eastern Road, Brighton BN2 5BE.

## Abstract

**Objectives:** To determine the association between frailty and medication-related harm requiring healthcare utilisation.

**Design:** Prospective observational cohort study.

**Setting:** 6 primary and 5 secondary care sites across South East England, September 2013 – November 2015.

**Participants:** 1280 participants, 65 years old or over, who were due for discharge from general medicine and older persons’ wards following an acute episode of care. Exclusion criteria were limited life expectancy, transfer to another hospital and consent not gained.

**Main outcome measures:** Medication-related harm requiring healthcare utilisation, including adverse drug reactions, non-adherence and medication error determined via triangulation of: patient/carer reports gathered through a structured telephone interview; primary care medical record review; and prospective consultant-led review of re-admission to recruiting hospital. Frailty was measured using a Frailty Index, developed using a standardised approach. Marginal estimates were obtained from logistic regression models to examine how probabilities of healthcare service use due to medication-related harm were associated with increasing polypharmacy and frailty.

**Results:** Healthcare utilisation due to medication-related harm was significantly associated with frailty (OR = 10.06, 95% CI 2.06 to 49.26, p = 0.004), independent of age, gender, and polypharmacy. With increasing polypharmacy, the need for healthcare use as a result of MRH increases from a probability of around 0.2 to 0.4. This is also the case for frailty.

**Conclusions:** Frailty is as important as polypharmacy in its association with MRH. Reducing the burden of frailty through an integrated health and social care approach, alongside strategies to reduce inappropriate polypharmacy, may reduce MRH related healthcare utilisation.

**Trial registration:** Approved by the National Research Ethics Service, East of England (REC Reference 13/EE/0075)

## Introduction

Frailty and medication-related harm (MRH) are two major challenges that increase the risk of poor outcomes such as hospital admission and death in older adults.(1) MRH includes harm arising from adverse drug reactions (ADRs), non-adherence and medication errors and affects one in three older adults following an acute hospital episode, costing approximately £400 million to the National Health Service (NHS).(2) The World Health Organisation (WHO) has challenged member states to reduce avoidable MRH by 50% by 2022.(3)

Frailty is defined as a state of increased vulnerability to suboptimal restoration of homoeostasis after a stressor event, and increases the risk of adverse outcomes(4). Frailty affects 14% of adults >50 years old in the United Kingdom, and the prevalence increases exponentially with age(5). In the United States it is reported to be increasing across younger adults. Whilst frailty lethality remains stable; the proportion of adults living with frailty is increasing.(6)

In a secondary analysis of inpatient hospital admission data (n=737) collected between 2012-13 in Ireland, the interaction between ADRs and frailty showed that frailty was more predictive of ADRs than the number of medicines alone. In this Irish study, frailty was identified using a frailty index, consisting of 34 items across a range of domains (including polypharmacy). A higher frailty index was associated with a greater likelihood of potentially inappropriate prescriptions and of experiencing at least one ADR during hospital admission. Patients taking more than six medicines were three times more likely to have at least one instance of potentially inappropriate prescriptions, but ADR occurrence was not associated with the number of medicines.(7)

Investigating the interaction between frailty and the broader context of MRH, beyond merely focusing only on ADRs, might influence the way in which healthcare professionals approach mitigating MRH. Approaches to reducing MRH in older adults, have traditionally focused on polypharmacy, medicines appropriateness and medicines reconciliation, but have thus far demonstrated limited impact on quality of life, clinical or economic outcomes.(8,9) The influence of frailty on MRH has been little explored, and crucially needs to be investigated independent of polypharmacy.

This study, therefore, aimed to determine whether frailty is independently associated with MRH requiring healthcare utilisation within a UK multicentre prospective cohort study (The Predicting RIsk of Medication-related harm in Elderly (PRIME) study).(2)

## Methods

### Study Participants

The data presented here relate to a sub-study of the PRIME study, the methods for which are described in detail in the published protocol(10). The PRIME study was approved by the National Research Ethics Service, East of England (REC Reference 13/EE/0075), and was funded by NIHR Research for Patient Benefit and Guy’s and St. Thomas’ Charity.

In summary, PRIME was a multicentre prospective cohort study of patients over the age of 65 years old. Patients from medical wards of five teaching hospitals in South England were invited to participate, immediately prior to hospital discharge. Those transferred to another acute healthcare setting (excluding transfer to intermediate care facilities), had a short life expectancy and were unlikely to survive to the end of the study eight week follow up period, or lacked capacity with no nominated consultee were excluded.

Baseline data included demographic, clinical, functional, psychological, and social data. MRH and associated healthcare utilisation within 8-weeks post-discharge were identified by senior pharmacists using three sources: (1) primary care records; (2) patient telephone interviews and (3) prospective review of hospital readmission in conjunction with the admitting medical consultant. MRH included ADRs, non-adherence and medication errors.

### Measures

#### Medication-related harm

MRH was classified as “doubtful”, “possible”, “probable” or “definite” and verified by an independent end-point committee, consisting of consultant geriatricians and a professor of clinical pharmacy. Events classified as ‘possible, probable and, definite’ were considered MRH.

#### Frailty

Frailty was measured in the PRIME study cohort through the development of a Frailty index (FI) using the approach described by Searle and colleagues(11); development of a FI was necessary because existing frailty measures e.g. Fried frailty phenotype(12) and the Clinical Frailty Scale(13) were not routinely recorded, and the electronic FI was developed for primary care datasets(14) and therefore not appropriate for this hospital cohort. The approach adopted was based on the premise that the more health-related deficits a person has, the more likely they are to be frail.(15) The number of deficits were counted and a ratio of the total number of deficits accumulated to the number of deficits considered calculated. Consistency has been demonstrated in this approach, even when the number and type of deficits differ(15–22); the frailer the person (that is, the higher the number of deficits), the more susceptible they are to adverse outcomes, including death(15).

Five criteria should be considered when developing a FI(11), four of which were appropriate for this study: 1. Variables must be deficits associated with health status; 2. The prevalence of the deficit must increase with age; 3. Deficits must not saturate too early; 4. Deficits must cover a range of systems. A fifth criteria was suggested (serial use of a single FI in the same population should include the same items each time) but was not applicable to our study where the FI could only be applied during the one episode of inpatient admission.

Finally, in the development of a FI there is no set number of deficits to be included however at least 30-40 deficits is recommended, and generally the more variables included the more precise is the index(11).

Following these standards, a FI was developed for the PRIME study cohort including 55 deficits from multiple domains (morbidity, cognition, mood, strength and mobility, nutrition, daily function) (Table 1). Furthermore, in accordance with the standardised approach, the relationship between frailty and mortality was determined with the aim of validating the FI developed in this study.(11) Kaplan-Meier plots were generated comparing survival in frail and non-frail patient using the established cut-off of 0.2, reflective of an individual approaching a frail state.(11,12,18,19) Validity of our 55-item FI was demonstrated; frail patients had significantly reduced 18-month survival. Median survival of frail patient was 14.2 months (95% CI 13.8-14.8) compared to non-frail patients 16.6 months (95% CI 16.3-16.9) (Log-Rank test p<0.001) (**Error! Reference source not found**.).

**Table 1.** Domains and deficits included in Frailty Index

| Domains | Variable | Deficit Coding |
| --- | --- | --- |
| Morbidity | *Charlson co-morbidities* |  |
|  | HIV/AIDS | Yes = 1, No = 0 |
|  | Connective tissue disorder | Yes = 1, No = 0 |
|  | Any dementia | Yes = 1, No = 0 |
|  | Any malignancy | Yes = 1, No = 0 |
|  | GI haematological | Yes = 1, No = 0 |
|  | Any liver disease | Yes = 1, No = 0 |
|  | CKD | Yes = 1, No = 0 |
|  | Diabetes | Yes = 1, No = 0 |
|  | Asthma/COPD | Yes = 1, No = 0 |
|  | Hemiplegia | Yes = 1, No = 0 |
|  | Previous stroke/TIA | Yes = 1, No = 0 |
|  | Peripheral vascular disease | Yes = 1, No = 0 |
|  | CHF | Yes = 1, No = 0 |
|  | Previous MI | Yes = 1, No = 0 |
|  | *Other diagnosis* |  |
|  | Glaucoma | Yes = 1, No = 0 |
|  | Hypertension | Yes = 1, No = 0 |
|  | IHD | Yes = 1, No = 0 |
|  | Osteoporosis | Yes = 1, No = 0 |
|  | Parkinsons | Yes = 1, No = 0 |
|  | Seizures | Yes = 1, No = 0 |
|  | Thyroid dysfunction | Yes = 1, No = 0 |
|  | Peripheral neuropathy | Yes = 1, No = 0 |
|  | Other cardiac disease | Yes = 1, No = 0 |
|  | Anaemia | Yes = 1, No = 0 |
|  | Hiatus hernia | Yes = 1, No = 0 |
|  | Gout | Yes = 1, No = 0 |
|  | TB | Yes = 1, No = 0 |
|  | VTE | Yes = 1, No = 0 |
|  | Atrial fibrillation | Yes = 1, No = 0 |
|  | Depression | Yes = 1, No = 0 |
|  | Hyperlipidaemia | Yes = 1, No = 0 |
|  | *Sensory* |  |
|  | Impaired hearing (and not corrected with hearing aid) | Yes = 1, No = 0 |
|  | Retinopathy | Yes = 1, No = 0 |
|  | Eyesight impaired (and not corrected with glasses) | Yes = 1, No = 0 |
|  | *Falls* |  |
|  | ≥2 falls in 12 months | Yes = 1, No = 0 |
|  | *Laboratory* |  |
|  | Hyponatraemia (Na < 135 mmol/l) | Yes = 1, No = 0 |
|  | Abnormal Hb: Female <121 g/l, Male <138 g/l | Yes = 1, No = 0 |
| Cognition | AMTS <8 | Yes = 1, No = 0 |
|  | Anxiety screen GAD-2 ≥3 | Yes = 1, No = 0 |
|  | Depression screen PHQ-2 ≥3 | Yes = 1, No = 0 |
| Strength | Grip strength: Female <14kg, Male <24kg<sup>#</sup> | Yes = 1, No = 0 |
| Function* | Help with stairs | Yes = 1, No = 0 |
|  | Help with mobility on level surfaces | Yes = 1, No = 0 |
|  | Help with transfers bed to chair and back | Yes = 1, No = 0 |
|  | Help with toilet use | Yes = 1, No = 0 |
|  | Help with bladder | Yes = 1, No = 0 |
|  | Help with bowels | Yes = 1, No = 0 |
|  | Help with dressing | Yes = 1, No = 0 |
|  | Help with grooming | Yes = 1, No = 0 |
|  | Help with bathing | Yes = 1, No = 0 |
|  | Help with feeding | Yes = 1, No = 0 |
| Nutrition | MUST ≥1 | Yes = 1, No = 0 |
|  | Albumin <35g/l | Yes = 1, No = 0 |
| Social | Has social care package (any frequency) | Yes = 1, No = 0 |
|  | Use of multi-compartment medicines compliance aid | Yes = 1, No = 0 |

<sup>#</sup> Median grip strength of study population stratified by gender used, to allow differentiation in a hospitalised and likely deconditioned population. 77.4% of females and 68.1% of males would be described as sarcopenic according to EWGSOP 2010(23) cut off of <20kg and <30kg, respectively
\*Barthel index; 1=dependent/needs help, 0=independent

### Analysis

To investigate the extent of how frailty influences healthcare utilisation following MRH, we developed a logistic regression model to estimate their association, considering concurrent influences of age, gender, and polypharmacy. We followed recommended procedures(24) for checking model misspecifications (Stata: linktest), need for non-linear terms (Stata: boxtid), multicollinearity (Stata: collin), Hosmer and Lemeshow’s goodness-of-fit test (Stata: lfit), and influential observations (Pearson residual, deviance residual and Pregibon leverage). We used average marginal effects at representative values(25) to illustrate how probabilities of healthcare service use for MRH vary with polypharmacy and frailty in each gender. Marginal estimation was carried out using the margins command that was introduced in Stata.(26) These estimates were bootstrapped 1000 times to obtain confidence intervals.

## Results

### Baseline demographics

The PRIME study recruited 1280 older adults at hospital discharge of which 17 (1.3%) died without follow-up, and 147 participants (11.5%) were lost to follow-up because no post-discharge information was available via readmission, GP records, or follow-up telephone call.(2) There was no clinically significant difference between those participants included or excluded from the final analysis (Table 2).

**Table 2.** Baseline sample characteristics of patient cohort

| Characteristic | Included patients†<br>(n=1116) | Excluded patients<br>(n=164) | p-value |
| --- | --- | --- | --- |
| Age, median (IQR), years | 81.9 (75.5-86.9) | 80.5 (74.7-86.2) | 0.123 |
| Gender, n (%) |  |  |  |
| Women | 652 (58.4) | 93 (56.7) | 0.673 |
| Men | 464 (41.6) | 71 (43.3) |  |
| Hospital stay, median (IQR), days | 7 (3-14) | 7 (3-13) | 0.595 |
| Number of Charlson Index comorbidities (%) |  |  |  |
| 0-1 | 541 (48.5) | 88 (53.7) |  |
| ≥2 | 575 (51.5) | 76 (46.3) | 0.242 |
| Selected comorbidities, n(%) |  |  |  |
| Hypertension | 611 (54.7) | 86 (52.4) | 0.615 |
| CLD | 326 (29.2) | 56 (34.1) | 0.202 |
| Atrial Fibrillation | 279 (25.0) | 43 (26.2) | 0.773 |
| Diabetes | 269 (24.1) | 31 (18.9) | 0.167 |
| IHD | 224 (20.1) | 38 (23.2) | 0.352 |
| CKD | 153 (13.7) | 21 (12.8) | 0.808 |
| CCF | 150 (13.4) | 20 (12.2) | 0.713 |
| Depression | 95 (8.5) | 12 (7.3) | 0.762 |
| Dementia | 51 (4.6) | 6 (3.7) | 0.839 |
| Charlson Index, median (IQR) | 2 (1-3) | 1 (1-3) | 0.087 |
| Bathel Score, median (IQR) | 17 (13-20) | 18 (14-20) | 0.035 |
| Hand grip strength*, median (IQR) | 18 (12-24) | 18 (12-26) | 0.345 |
| Falls* (2 or more in last year), n (%) | 401 (36.4) | 57 (35.2) | 0.794 |
| Number of discharge medicines, median (IQR) | 9 (7-12) | 9 (6-12) | 0.393 |
| Multicompartment compliance aid, n (%) | 371 (33.2) | 43 (26.2) | 0.074 |
| Discharge to care home, n (%) | 30 (2.7) | 8 (4.9) | 0.136 |
| Living alone after discharge, n (%) | 551 (49.4) | 80 (48.8) | >0.999 |

IQR: Interquartile range; CLD: chronic lung disease; IHD: ischaemic heart disease; CKD: chronic kidney disease; CCF: congestive cardiac failure.
†ten patients were included following readmission which was not associated with MRH, for whom GP records were not available and were uncontactable at 8-weeks (median follow-up 29 days after recruitment). \*missing data: hand grip strength, n=164; falls, n=15.
Mann-Whitney U test for continuous variables and Fisher's Exact test for categorical variables

The median age of the PRIME study cohort was 82 years old (IQR 75.5-86.9 years) with more female participants (58%). The study population had a high level of multi-morbidity (52% had a Charlson Comorbidity Index of ≥2), with cardiac and respiratory conditions, and diabetes dominating. The level of multimorbidity was reflected in the number of discharge medicines (median number of discharge drugs=9, IQR 7-12, range 0-27) and one third of study participants (33.2%) used a multi-compartment medicines compliance aid to support medicines use. There was a median of two drug changes per participant between admission and discharge. Participants had an average length of stay of one week (median 7, IQR 3-14 days).

Median hand grip strength for the study population was 18kg (IQR 12-24kg) and when differentiated by gender 77.4% of females and 68.1% of males would be described as sarcopenic(23), with median hand grip strength of 14kg (IQR 10-18kg) and 24kg (IQR 19-31kg) respectively. Using a clinically relevant cut-point of 0.2(15), 446 patients (40%) were frail with a FI range from 0-0.44.

Out of 1112 participants (4 were excluded due to incomplete MRH outcome data), 413 (37%) participants experienced MRH in the 8-week follow up period and 52% of the MRH events were potentially preventable. Healthcare utilisation secondary to MRH was required in 328 (29%) participants, equating to 96 hospital admissions and 316 GP consultations. Further details, including types of MRH and medications involved have been published elsewhere.(2)

Those requiring MRH-related healthcare use tended to be older, female, taking more medicines and frailer (Table 3).

**Table 3.**
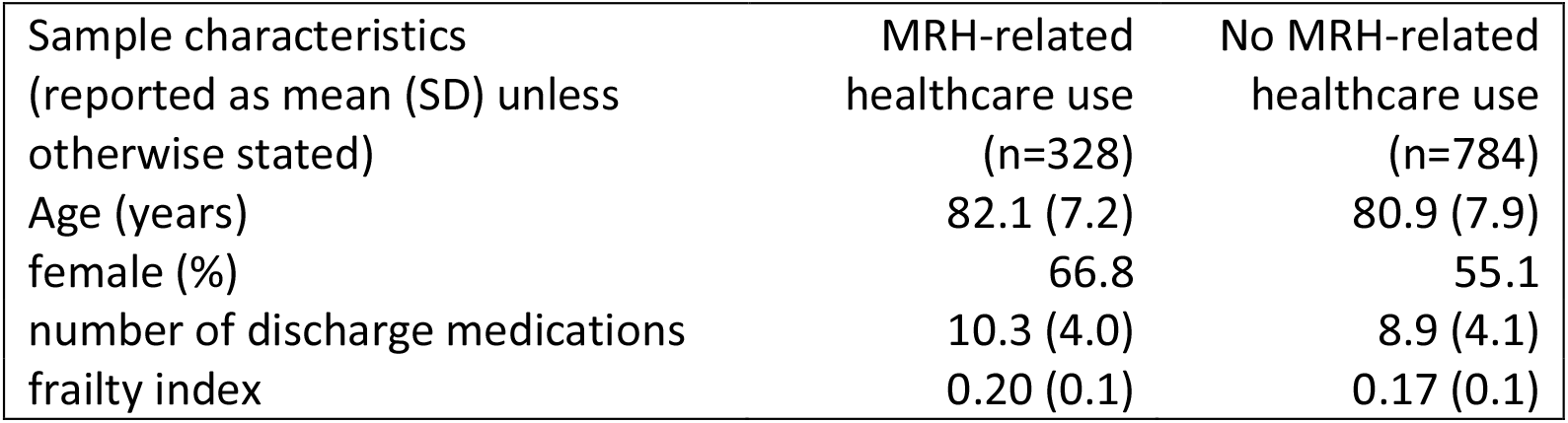
Sample description

| Sample characteristics<br>(reported as mean (SD) unless<br>otherwise stated) | MRH-related<br>healthcare use<br>(n=328) | No MRH-related<br>healthcare use<br>(n=784) |
| --- | --- | --- |
| Age (years) | 82.1 (7.2) | 80.9 (7.9) |
| female (%) | 66.8 | 55.1 |
| number of discharge medications | 10.3 (4.0) | 8.9 (4.1) |
| frailty index | 0.20 (0.1) | 0.17 (0.1) |

### Frailty associated with MRH

Healthcare utilisation due to MRH showed statistically significant associations with frailty (OR = 10.06, 95% CI 2.06 – 49.26, p = 0.004), taking into account concurrent influence of age, gender, and polypharmacy (Table 4). The odds of healthcare service utilisation for MRH were increased with polypharmacy and frailty but were lower in males. These odds were not associated with age, possibly because this study’s population was an older cohort.

**Table 4.**
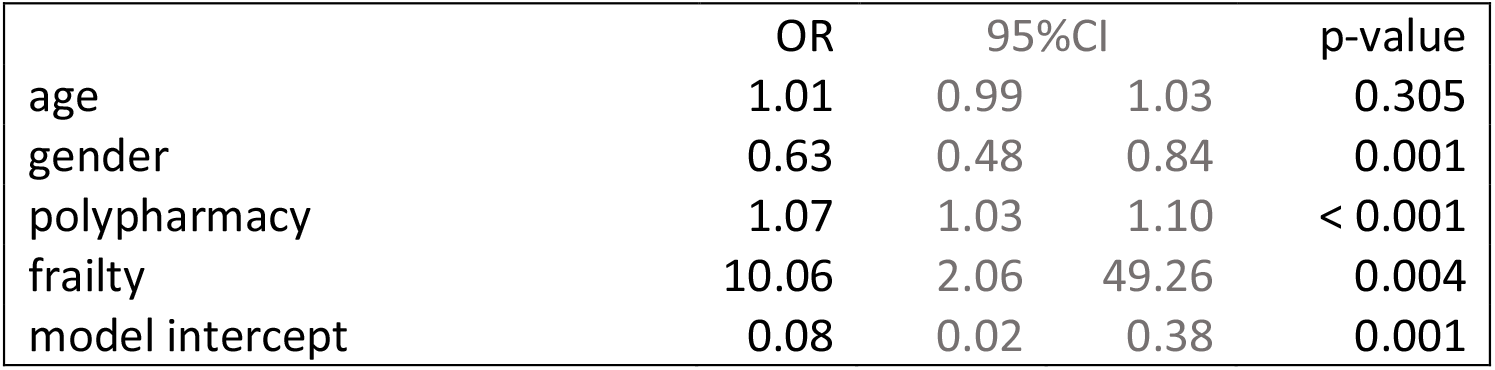
Odds ratios (ORs) from logistic regression model (dependent variable: healthcare service use due to medication-related harm)

Female gender tended to be associated with elevated risks for MRH healthcare utilisation, particularly when the number of medications was in the range of 7 – 13 (**Error! Reference source not found**.) and the frailty index was in the range of 0.15 – 0.25 (**Error! Reference source not found**.). Detailed estimates and bootstrapped confidence intervals are reported in the online supplement. With increasing polypharmacy, the need for healthcare utilisation as a result of MRH increases from a probability of around 0.2 to 0.4. This is also the case for frailty. Frailty appears to matter as much as polypharmacy in its association with MRH.

### Sensitivity analysis

Reflective of clinical practice, and in line with similar research in this area, we also modelled frailty as a binary variable, using 0.2 as the cut-point. Using this approach resulted in a marked difference in the association between frailty and MRH healthcare use, when compared to modelling frailty as a continuous variable: frailty binary OR = 1.37 (95%CI: 1.04 – 1.81, p = 0.027) (online supplement), frailty continuous OR = 10.06 (95%CI: 2.06 – 49.26, p = 0.004) (Table 4).

## Discussion

### Statement of principal findings

This study demonstrated that frailty is a predictor of healthcare utilisation secondary to MRH in the 8 weeks post-hospital discharge, independent of polypharmacy. The need for healthcare intervention reflects both the severity of MRH, and the vulnerability of older adults living with frailty to the harmful effects associated with medicines. Furthermore, our findings suggest that frailty is an important driver of MRH, in addition to polypharmacy, which should prompt review of current approaches to tackling the global challenge of MRH in older adults.

### Strengths and weaknesses of the study

The major strengths of this study were the large number of participants, its prospective multicentre design, a robust multistage, multidisciplinary process for verifying MRH, and a validated frailty index. However, an important limitation was measuring frailty using an index that has only been internally validated, and this may impact on the generalisability of our findings. Efforts were made to minimise this limitation by following the standardised approach to developing a Frailty Index, and through inclusion of a larger number of variables than the recommended minimum: an approach which has been reported to improve precision.(11)

### Strengths and weaknesses in relation to other studies

Traditionally, the number of medicines has been used as a trigger for medicines review in clinical practice(27), and deprescribing interventions, including the application of medicines appropriateness criteria as recommended by NHS England(28). Whilst these interventions have reduced the number of medicines prescribed, and increased their appropriateness, their long-term impact on clinical outcomes has been limited.(8,9) Polypharmacy is logically and evidently a dominant driver of MRH, but our study demonstrates that frailty is similarly important. On-going research to understand the risk factors associated with MRH has illustrated the importance of not only biological or medicines-related variables but also the impact of psychological and social variables on MRH. “Living alone”, a social determinant, was identified as an important predictor of MRH in our development of an MRH risk-prediction tool.(29) Hence MRH should be viewed through a holistic bio-psychosocial lens.(30) The complexity of the relationship between MRH, frailty and polypharmacy is also seen in clinical practice; not all patients with multiple comorbidities and polypharmacy will experience MRH, similarly patients with few medicines, but multiple social and psychological challenges may experience harm. As with frailty and other geriatric syndromes, viewing MRH from this perspective recognises the multifactorial nature of the problem, and the need for a more sophisticated and holistic approach to its mitigation.

Our results resonate with the findings of Lattanzio *et al*(31) who investigated the association between Geriatric Conditions and ADRs in 506 hospitalised older adults. Adverse drug reactions were experienced by 11.5% of inpatients (mean age 80.1 years, SD 6.0 years; 54.3% female; mean number of medicines 10.6, SD 5.5). Whilst there was no association between individual Geriatric Conditions and ADRs, the combined variable of history of falls and dependency in at least one ADL was significantly correlated with ADRs (OR 2.18, 95% CI 1.13-4.19) and upon multivariable analysis ADR was independent of the number of medicines used. It may be argued that whilst singular geriatric conditions are markers of frailty, individually they do not reflect the global loss of homeostatic reserve seen in frailty that limits an individual’s ability to withstand a situational challenge presented by a medicine. The presence of more than one geriatric condition, in particular, dependency in ADLs in the context of an acute episode of care may reflect a more globally compromised functional reserve system, thus making these individuals more vulnerable to an ADR.

Our study identified an elevated risk of MRH requiring healthcare utilisation for females. This may partly be explained by, globally, older females have increased difficulties in performing instrumental activities of daily living, which includes medication-taking, compared to men.(32) Therefore, as with frailty, future interventions must incorporate the social aspect of MRH risk. In drawing this conclusion, the increased likelihood that females seek help with their health (33) and so have more opportunity for MRH requiring healthcare to be identified and recorded should be considered.

### Meaning of the study

We found that frailty substantially increases the risk of older adults experiencing MRH that requires further management. It would seem sensible and imperative then to target primary care interventions at those with the highest levels of frailty. Although, frail older adults may already benefit from geriatrician intervention, where medication review forms part of the Comprehensive Geriatric Assessment, our research suggests that more needs to be done, for example community based follow up, responsive to the dynamic nature of frailty. Future strategies in primary care should also adopt a more proactive approach to mitigating MRH by focusing on individuals who are approaching frailty or living with mild frailty, and not yet known to specialist geriatric services. The increasing prevalence of frailty in younger years(6), and the Health Secretary’s ambition to add “years to life, and life to years” should motivate this change.

The findings of our study also stress the importance of avoiding the dichotomisation of continuous variables due to the risk of loss of valuable data(34). Dichotomisation of frailty in this study resulted in loss of information about its influence on MRH requiring healthcare, which when applied in practice may incorrectly influence resource allocation. Furthermore, as highlighted by the proposal to apply frailty scales to determine the access of older people to healthcare during the COVID-19 pandemic, we are reminded that frailty is not a binary state, but a bi-directional continuum.(35)

### Unanswered questions and future research

Frailty is a risk factor for MRH requiring healthcare utilisation, independent of polypharmacy. Further exploration of the interactions between these two geriatric syndromes is required to inform future health and social care policy and practice to mitigating MRH. A holistic, individualised approach to reducing the burden of frailty, alongside strategies to reduce inappropriate polypharmacy, may reduce the incidence of MRH and subsequent healthcare utilisation.

**Figure 1.**
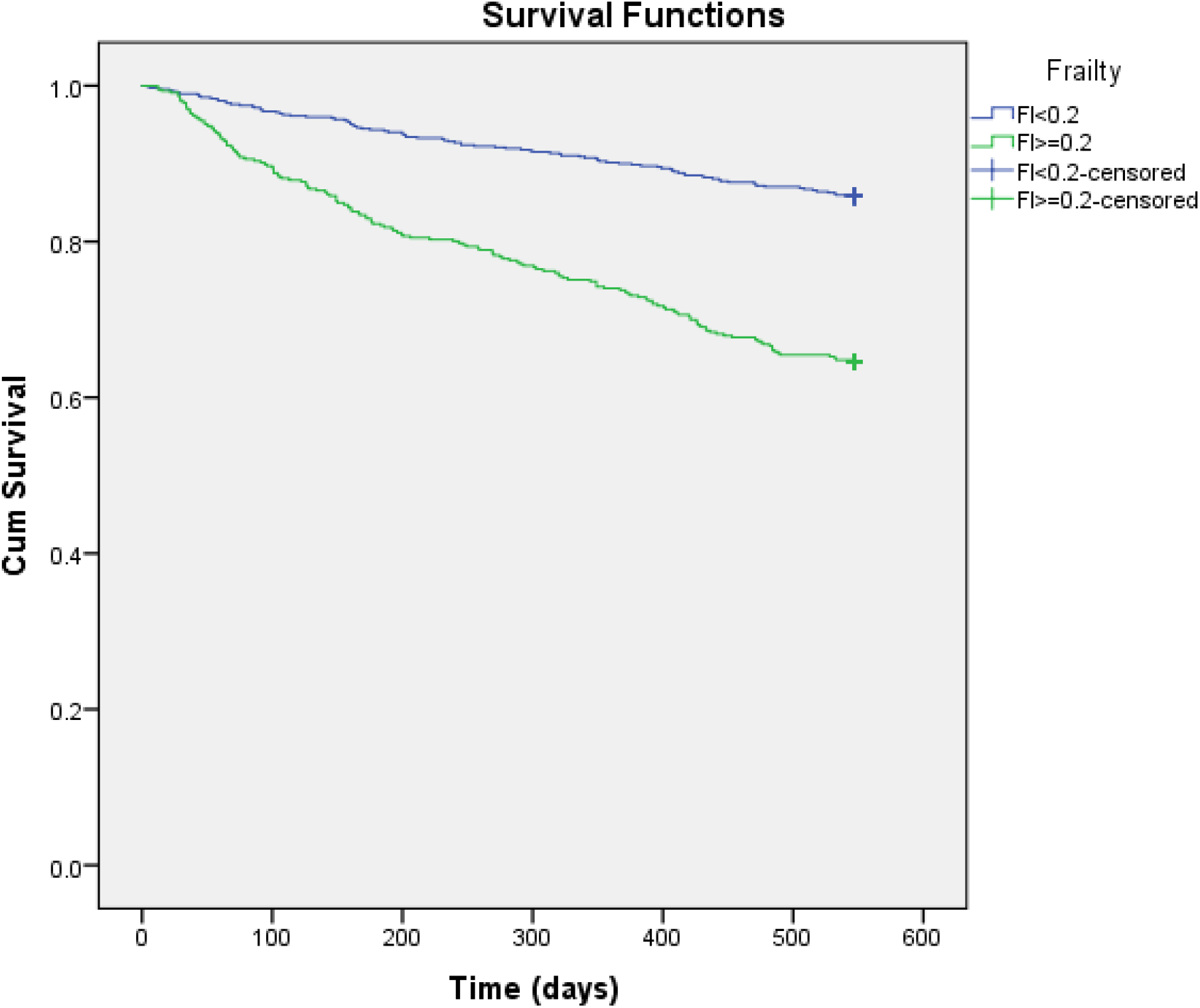
Kaplan-Meier plot comparing 18-month survival of frail (FI ≥0.2) and non-frail (FI < 0.2) patients.

**Figure 2.**
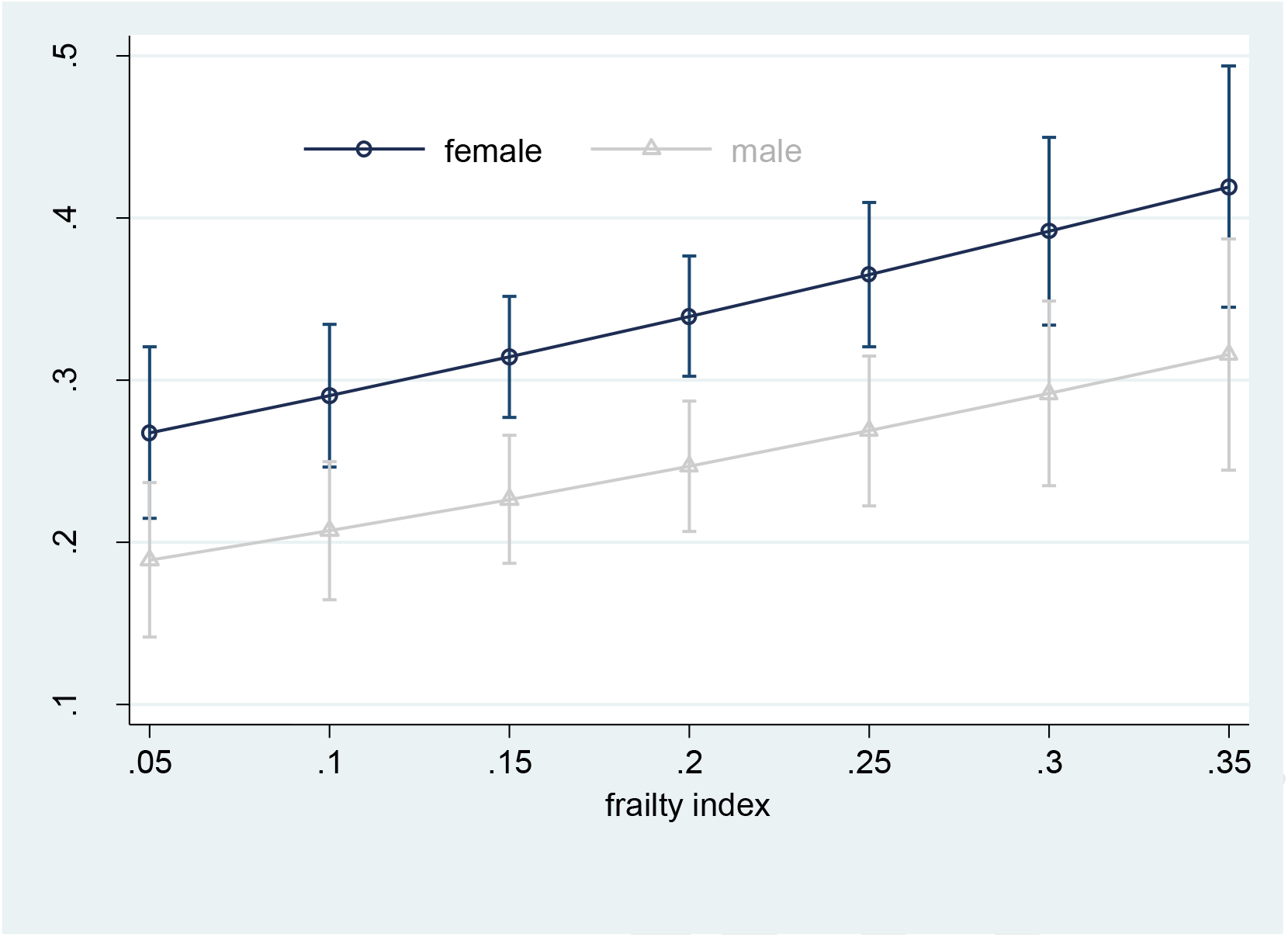
Average marginal estimates (95% confidence intervals) of probabilities of healthcare service use due to medication-related harm at representative values of frailty index

**Figure 3.**
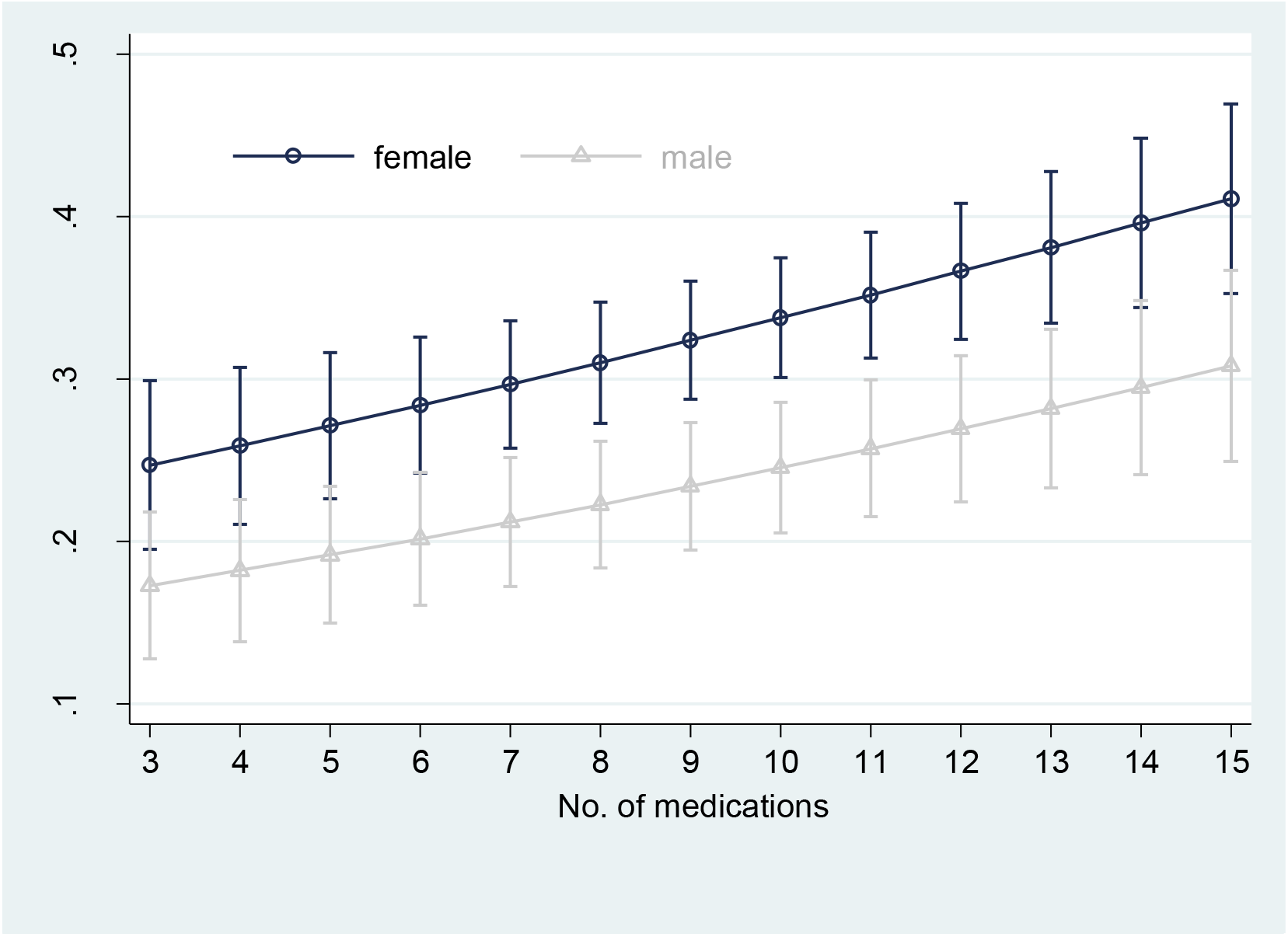
Average marginal estimates (95% confidence intervals) of probabilities of healthcare service use due to medication-related harm at representative values of polypharmacy

## Data Availability

All data relevant to the study are included in the article or uploaded as supplementary information. Further relevant anonymised patient level data is available on reasonable request.

## Contributors

JMS, JGD, RS, CR, KA conceived the PRIME Study. JGD, RS, KA, CR were co-applicants for the study grant. JMS, NP, JGD, RS, CR, KA designed the study. Data analysis was performed by JMS, NP, KCC, with KCC providing statistical expertise. All authors were involved in preparing the manuscript. All authors reviewed and approved the final manuscript. CR is the guarantor and as the corresponding author attests that all listed authors meet authorship criteria and that no others meeting the criteria have been omitted.

## Copyright

Professor C Rajkumar (corresponding author) has the right to grant on behalf of all authors and does grant on behalf of all authors, an exclusive licence (or non exclusive for government employees) on a worldwide basis to the BMJ Publishing Group Ltd to permit this article (if accepted) to be published in BMJ editions and any other BMJPGL products and sublicences such use and exploit all subsidiary rights, as set out in our licence.

## Transparency declaration

JMS and CR affirm that the manuscript is an honest, accurate, and transparent account of the study being reported; that no important aspects of the study have been omitted; and that any discrepancies from the study as planned (and, if relevant, registered) have been explained.

## Statement of independence of researchers from study funders

all researchers were independent from study funders.

## Patient and Public Involvement

In June 2012, at St. Thomas’ Hospital, London, JMS led a focus group of six patients with chronic disease, taking multiple medications, who had a recent acute hospital admission. In this meeting, JMS explored patients’ experience of medication problems and contributing factors to help guide relevant data collection. Patients provided feedback on the patient information sheet, and confirmed the importance of this research for older people. There was no patient involvement in data collection, analysis or interpretation. The research will be presented to the public at regional educational events.

## Dissemination Declaration

Dissemination of findings to study participants in not possible, but the wider findings of the PRIME study have been shared with involved healthcare organisations and the general public through media outlets, including local television and national and international newspapers.

## Patient consent for publication

not required

