## Supplementary File for "Frailty is a predictor of medication-related harm requiring healthcare utilisation: a multicentre prospective cohort study"

#### Model diagnostics

A logistic regression model was used to estimate the association between healthcare service use (mrh9) and frailty index (frp), taking into account the concurrent influence of age (age), gender (male), and polypharmacy (poly).

We followed recommended procedures for examining model diagnostics (UCLA Statistical Consulting Group: https://stats.idre.ucla.edu/stata/webbooks/logistic/chapter3/lesson-3-logistic-regression-diagnostics-2/).

##### Collinearity

For multicollinearity, variance inflation factor (VIF) of 10 or greater (or tolerance of 0.1 or less) would be a cause for concern. The frailty index had the largest VIF (1.25). This suggests that the large standard error (Table X in main text) is due to collinearity. However, the magnitude of VIF did not raise major concerns with multicollinearity among the independent variables.

| Model diagnostics | Stata command |
|---|---|
| multicollinearity | collin age male poly frp if incl==1 |

Table S1. Collinearity diagnostics

| | VIF | Tolerance |
|---|---|---|
| age | 1.15 | 0.8694 |
| male | 1.03 | 0.9717 |
| poly | 1.13 | 0.8859 |
| frp | 1.25 | 0.8021 |

PREPRINT

#### Specification errors

To see if the model is properly specified, we examined the results of the linktest. The variable **_hat** should be a statistically significant predictor, since it is the predicted value from the model. The variable **_hatsq** did not attain statistical significance, indicative of the absence of model specification errors (e.g. no relevant variable omitted).

| Model diagnostics | Stata command |
|---|---|
| Logistic regression | quietly logit mrh9 age i.male poly frp if incl==1 |
| Model misspecification | linktest |

Table S2. Model diagnostics (linktest) for specification errors

| | p-value |
|---|---|
| _hat | 0.009 |
| _hatsq | 0.829 |
| _cons | 0.893 |

#### Non-linearity

The Box-Tidwell model tests the null hypothesis that the change of a dependent variable on a predictor is linear (i.e. p1 = 1). None of the terms attained statistical significance. This suggests that linearity assumptions of our logistic regression model are tenable.

| Model diagnostics | Stata command |
|---|---|
| non-linearity | boxtid logit mrh9 age male poly frp if incl==1 |

Table S3. Non-linearity diagnostics

| | p1 value | Non-linear deviation | p-value |
|---|---|---|---|
| age | -22.743 | 3.592 | 0.058 |
| poly | 0.227 | 1.737 | 0.188 |
| frp | 2.235 | 1.150 | 0.283 |

#### Goodnes-of-fit

The Hosmer and Lemeshow’s goodness-of-fit test yielded a large p-value (i.e. not statistically significant), indicative of a good match between the predicted and observed frequencies.

| Model diagnostics | Stata command |
|---|---|
| Logistic regression | quietly logit mrh9 age i.male poly frp if incl==1 |
| Hosmer and Lemeshow’s goodness-of-fit | lfit |

Pearson chi square (df=1107) = 1113.54, p = 0.4392

PREPRINT

#### Prediction errors

To look for observations with unusually large prediction errors, we inspected Pearson residual (standardized difference between the observed frequency and the predicted frequency) and deviance residual (disagreement between the maxima of the observed and the fitted log likelihood functions). To look for observations which might have larger than usual impact on model results, we inspected Pregibon leverage values.

| Influential observations | Stata command |
|---|---|
| Pearson residual | gen id=_n<br>quietly logit mrh9 age i.male poly frp if incl==1<br>predict p<br><br>predict zres, rstand<br>scatter zres p, yline(0) mlab(id) mlabsize(1.4) msize(0.5) |
| deviance residual | predict dvr, dev<br>scatter dvr p, yline(0) mlab(id) mlabsize(1.4) msize(0.5) |
| Pregibon leverage | predict lev, hat<br>scatter lev p, yline(0) mlab(id) mlabsize(1.4) msize(0.5) |

PRINT

Observation ID 226, 900 and 206 had slightly larger than usual Pearson residuals.

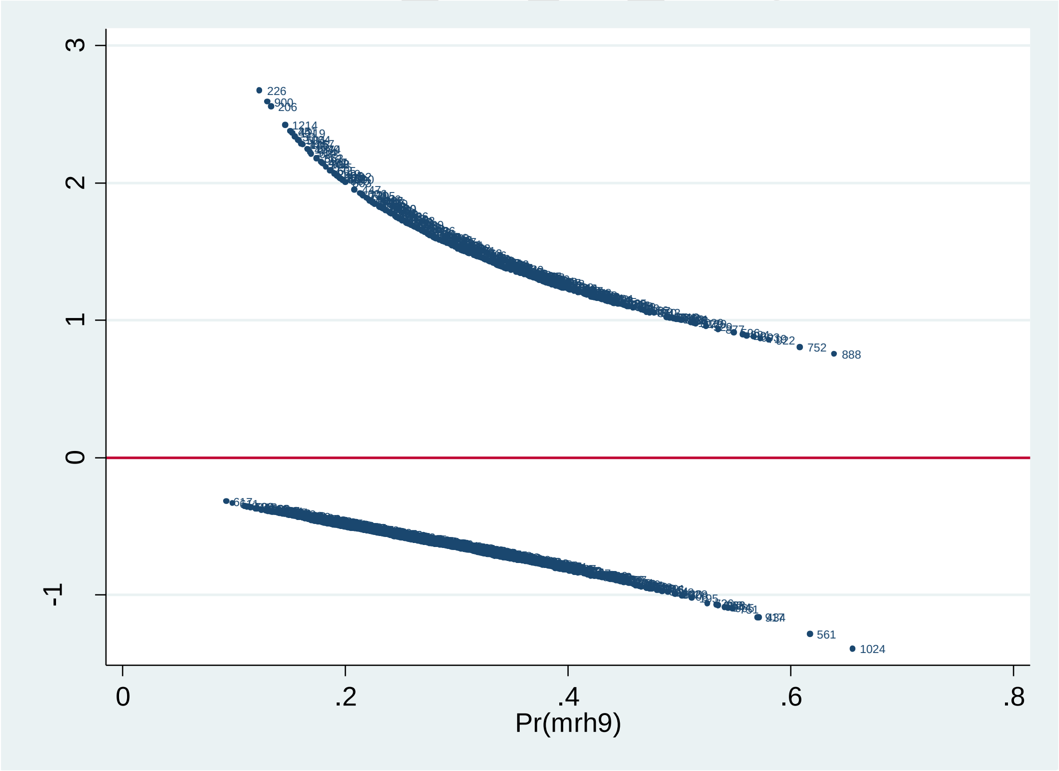

Figure S1 Pearson residual across predicted probabilities of healthcare service use due to medication-related harm

Observation ID 226, 900 and 206 had slightly larger than usual deviance residuals.

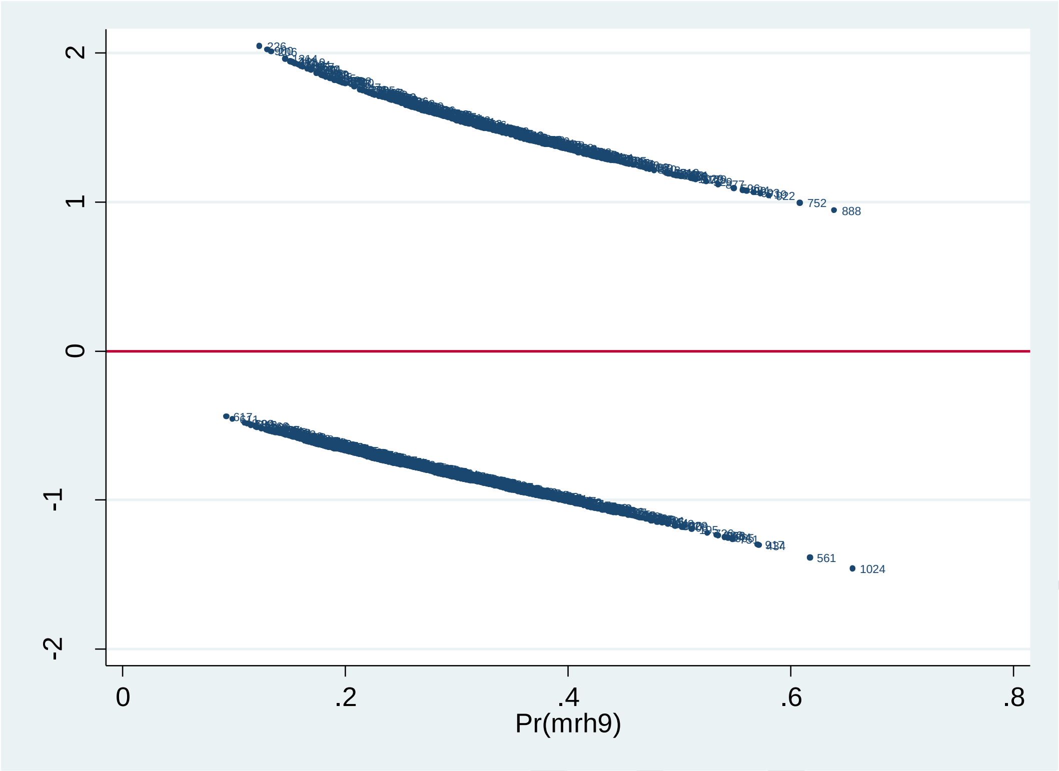

Figure S2 Deviance residual across predicted probabilities of healthcare service use due to medication-related harm

PREPRINT

Observation ID 726, 561, 917, and 1024 had slightly larger than usual leverage.

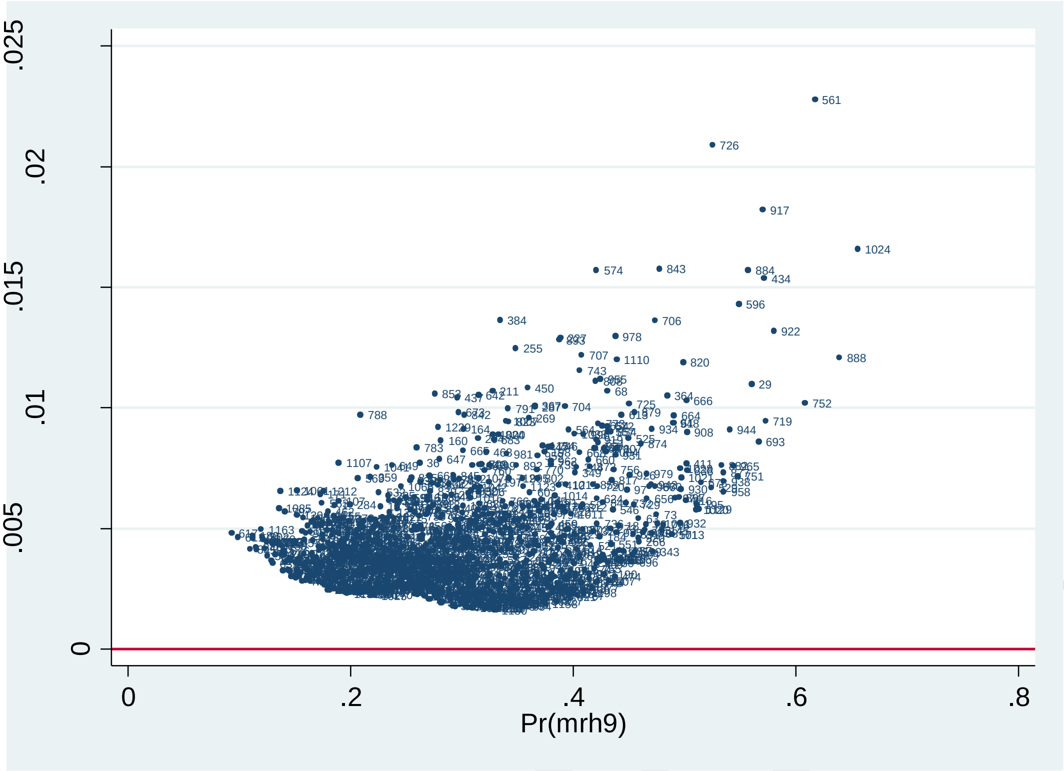

Figure S3 Pregibon leverage across predicted probabilities of healthcare service use due to medication-related harm

#### Diagnostics summary

Model diagnostics raised no major concern with model specifications, model assumptions, and model fit. Residual and influence statistics flagged a handful of observations that were slightly less typical or might have had a slightly larger than usual impact on model results. We conducted sensitivity analysis that excluded these observations. The model results gave almost identical estimates as the original analysis as described below.

PREPRINT

### Model results

#### Logistic regression models

We conducted sensitivity analysis that removed observation ID 226, 900, 206, 726, 561, 917, 1024 (Table S4). The model results gave almost identical estimates as the original analysis (Table S5, and Table 4 in main text).

Table S4. Odds ratios (ORs) from logistic regression model (dependent variable: healthcare service use due to medication-related harm)

| | OR | 95%CI | | p-value |
|---|---|---|---|---|
| age | 1.01 | 0.99 | 1.03 | 0.257 |
| gender | 0.62 | 0.47 | 0.82 | 0.001 |
| polypharmacy | 1.08 | 1.04 | 1.12 | < 0.001 |
| frailty | 11.95 | 2.41 | 59.15 | 0.002 |
| model intercept | 0.06 | 0.01 | 0.30 | 0.001 |

Table 4 in main text (inserted here for ease of reference)

| | OR | 95%CI | | p-value |
|---|---|---|---|---|
| age | 1.01 | 0.99 | 1.03 | 0.305 |
| gender | 0.63 | 0.48 | 0.84 | 0.001 |
| polypharmacy | 1.07 | 1.03 | 1.10 | < 0.001 |
| frailty | 10.06 | 2.06 | 49.26 | 0.004 |
| model intercept | 0.08 | 0.02 | 0.38 | 0.001 |

PREPRINT

#### Average marginal estimates – frailty

Table S8. Probabilities of MRH (average marginal estimates) by frailty levels and gender

| frailty index | female | male | gender difference | Lower CI (difference) | Upper CI (difference) |
|---|---|---|---|---|---|
| .05 | .27 | .19 | -.08 | -.13 | -.03 |
| .10 | .29 | .21 | -.08 | -.13 | -.03 |
| .15 | .31 | .23 | -.09 | -.14 | -.03 |
| .20 | .34 | .25 | -.09 | -.15 | -.04 |
| .25 | .37 | .27 | -.10 | -.16 | -.04 |
| .30 | .39 | .29 | -.10 | -.16 | -.04 |
| .35 | .42 | .32 | -.10 | -.17 | -.04 |

CI: bootstrapped confidence intervals

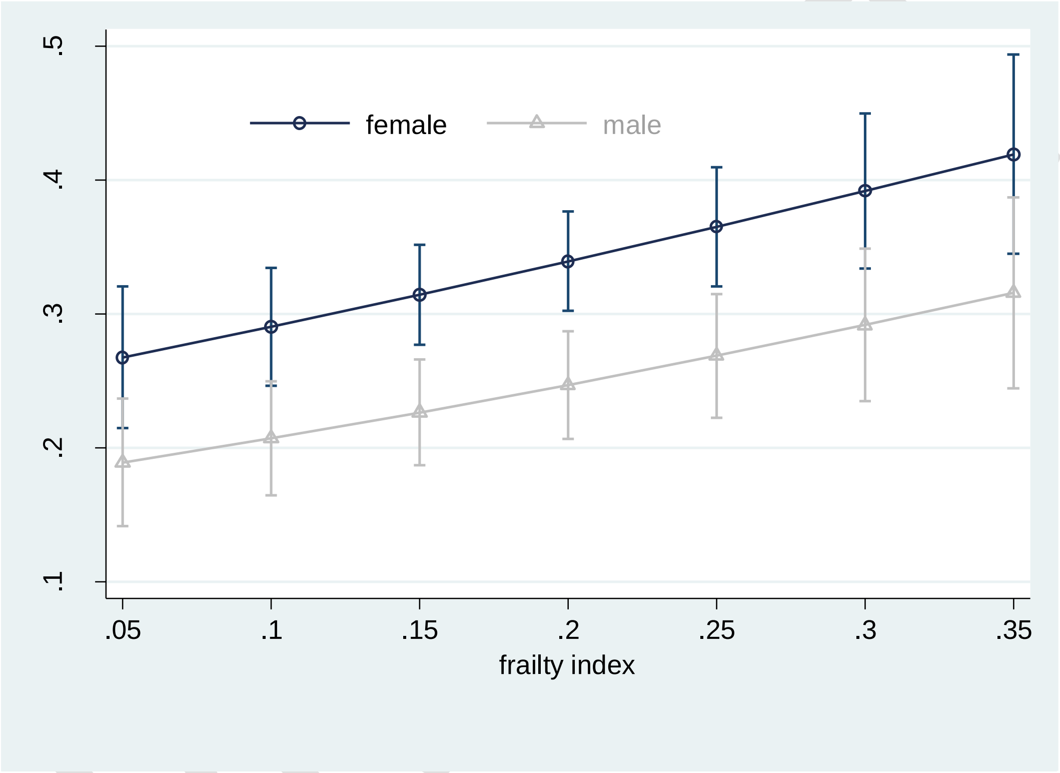

Figure 2 in main text (inserted here for ease of reference)

Average marginal estimates (95% confidence intervals) of probabilities of healthcare service use due to medication-related harm at representative values of frailty index

```
* # model 1 bootstrap margins for frailty
* #########################
qui logit mrh9 age i.male poly frp if incl==1
keep if e(sample)

mat shelf1 = (r(f1), r(f2), r(f3), r(f4), r(f5), r(f6), r(f7), r(m1),
r(m2), r(m3), r(m4), r(m5), r(m6), r(m7), r(d1), r(d2), r(d3),
r(d4), r(d5), r(d6), r(d7))

capture program drop pacman
prog define pacman, rclass
preserve
drop if incl==0

qui logit mrh9 age i.male poly frp
margins male, at(frp=(0.05(0.05)0.35)) post
mat box1 = e(b)
sca x1 = box1[1,1]
sca y1 = box1[1,2]
sca x2 = box1[1,3]
sca y2 = box1[1,4]
sca x3 = box1[1,5]
sca y3 = box1[1,6]
sca x4 = box1[1,7]
sca y4 = box1[1,8]
sca x5 = box1[1,9]
sca y5 = box1[1,10]
sca x6 = box1[1,11]
sca y6 = box1[1,12]
sca x7 = box1[1,13]
sca y7 = box1[1,14]

qui logit mrh9 age i.male poly frp
margins, dydx(male) at(frp=(0.05(0.05)0.35)) post
mat box2 = e(b)
sca z1 = box2[1,8]
sca z2 = box2[1,9]
sca z3 = box2[1,10]
sca z4 = box2[1,11]
sca z5 = box2[1,12]
sca z6 = box2[1,13]
sca z7 = box2[1,14]

return sca f1 = x1
return sca f2 = x2
return sca f3 = x3
return sca f4 = x4
return sca f5 = x5
return sca f6 = x6
return sca f7 = x7

return sca m1 = y1
return sca m2 = y2
return sca m3 = y3
return sca m4 = y4
return sca m5 = y5
return sca m6 = y6
return sca m7 = y7

return sca d1 = z1
return sca d2 = z2
return sca d3 = z3
return sca d4 = z4
return sca d5 = z5
return sca d6 = z6
return sca d7 = z7

restore
end

bootstrap ///
f1=r(f1) f2=r(f2) f3=r(f3) f4=r(f4) f5=r(f5) f6=r(f6) f7=r(f7)
///
m1=r(m1) m2=r(m2) m3=r(m3) m4=r(m4) m5=r(m5)
m6=r(m6) m7=r(m7)   ///
d1=r(d1) d2=r(d2) d3=r(d3) d4=r(d4) d5=r(d5) d6=r(d6)
d7=r(d7) ,            ///
reps(1000) seed(25091936): pacman

estat bootstrap, all

translate @Results "$taxi/mrhf boot1.txt", replace
```

PREPRINT

#### Average marginal estimates – number of medicines

Table S8. Probabilities of MRH (average marginal estimates) by number of medications and gender

| no. of medications | female | male | gender difference | Lower CI (difference) | Upper CI (difference) |
|---|---|---|---|---|---|
| 3 | .25 | .17 | -.07 | -.12 | -.03 |
| 4 | .26 | .18 | -.08 | -.13 | -.03 |
| 5 | .27 | .19 | -.08 | -.13 | -.03 |
| 6 | .28 | .20 | -.08 | -.13 | -.03 |
| 7 | .30 | .21 | -.08 | -.14 | -.03 |
| 8 | .31 | .22 | -.09 | -.14 | -.03 |
| 9 | .32 | .23 | -.09 | -.15 | -.03 |
| 10 | .34 | .25 | -.09 | -.15 | -.03 |
| 11 | .35 | .26 | -.09 | -.15 | -.04 |
| 12 | .37 | .27 | -.10 | -.16 | -.04 |
| 13 | .38 | .28 | -.10 | -.16 | -.04 |
| 14 | .40 | .29 | -.10 | -.16 | -.04 |
| 15 | .41 | .31 | -.10 | -.17 | -.04 |

CI: bootstrapped confidence intervals

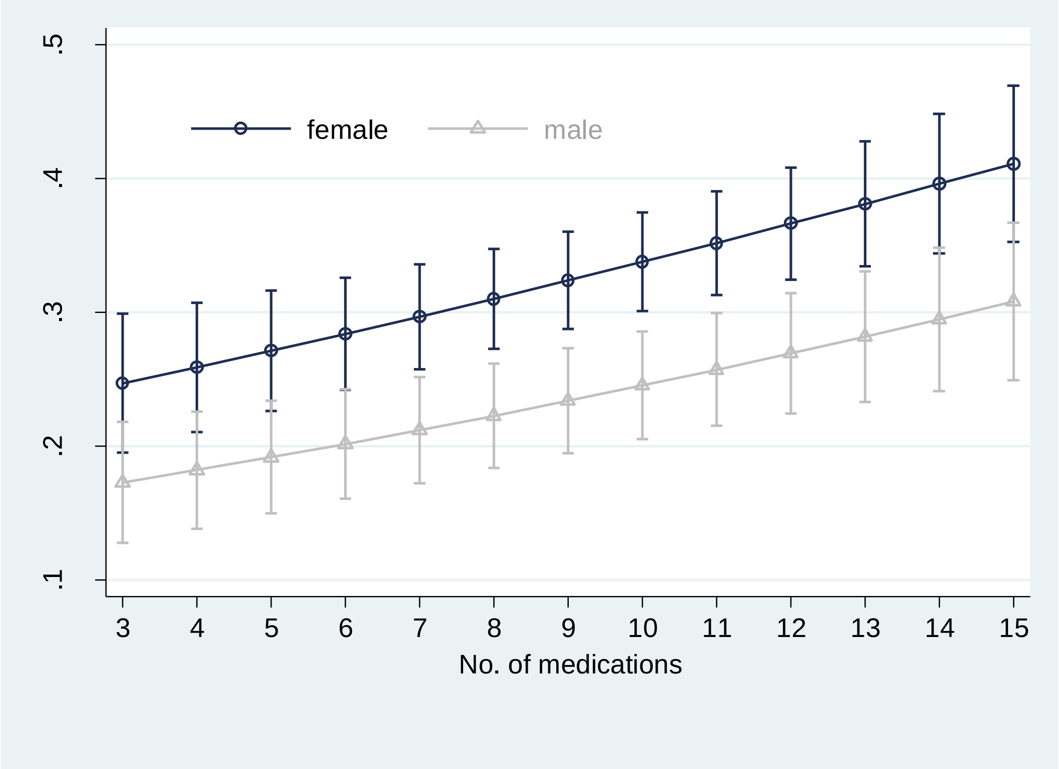

Figure 3 in main text (inserted here for ease of reference)

Average marginal estimates (95% confidence intervals) of probabilities of healthcare service use due to medication-related harm at representative values of polypharmacy

```
* # model bootstrap margins for polypharmacy
* ########################
qui logit mrh9 age i.male poly frp if incl==1
keep if e(sample)

mat shelf2 = ///
(r(f1), r(f2), r(f3), r(f4), r(f5), r(f6), r(f7) ,                ///
r(f8), r(f9), r(f10), r(f11), r(f12), r(f13) ,                 ///
r(m1), r(m2), r(m3), r(m4), r(m5), r(m6), r(m7) ,   ///
r(m8), r(m9), r(m10), r(m11), r(m12), r(m13) ,     ///
r(d1), r(d2), r(d3), r(d4), r(d5), r(d6), r(d7)) ,        ///
r(d8), r(d9), r(d10), r(d11), r(d12), r(d13)

capture program drop pacman
prog define pacman, rclass
preserve
drop if incl==0

qui logit mrh9 age i.male poly frp
margins male, at(poly=(3(1)15)) post
mat box1 = e(b)
sca x1 = box1[1,1]
sca y1 = box1[1,2]
sca x2 = box1[1,3]
sca y2 = box1[1,4]
sca x3 = box1[1,5]
sca y3 = box1[1,6]
sca x4 = box1[1,7]
sca y4 = box1[1,8]
sca x5 = box1[1,9]
sca y5 = box1[1,10]
sca x6 = box1[1,11]
sca y6 = box1[1,12]
sca x7 = box1[1,13]
sca y7 = box1[1,14]
sca x8 = box1[1,15]
sca y8 = box1[1,16]
sca x9 = box1[1,17]
sca y9 = box1[1,18]
sca x10 = box1[1,19]
sca y10 = box1[1,20]
sca x11 = box1[1,21]
sca y11 = box1[1,22]
sca x12 = box1[1,23]
sca y12 = box1[1,24]
sca x13 = box1[1,25]
sca y13 = box1[1,26]

qui logit mrh9 age i.male poly frp
margins, dydx(male) at(poly=(3(1)15)) post
mat box2 = e(b)
sca z1 = box2[1,14]
sca z2 = box2[1,15]
sca z3 = box2[1,16]
sca z4 = box2[1,17]
sca z5 = box2[1,18]
sca z6 = box2[1,19]
sca z7 = box2[1,20]
sca z8 = box2[1,21]
sca z9 = box2[1,22]
sca z10 = box2[1,23]
sca z11 = box2[1,24]
sca z12 = box2[1,25]
sca z13 = box2[1,26]
```

```
return sca f1 = x1
return sca f2 = x2
return sca f3 = x3
return sca f4 = x4
return sca f5 = x5
return sca f6 = x6
return sca f7 = x7
return sca f8 = x8
return sca f9 = x9
return sca f10 = x10
return sca f11 = x11
return sca f12 = x12
return sca f13 = x13

return sca m1 = y1
return sca m2 = y2
return sca m3 = y3
return sca m4 = y4
return sca m5 = y5
return sca m6 = y6
return sca m7 = y7
return sca m8 = y8
return sca m9 = y9
return sca m10 = y10
return sca m11 = y11
return sca m12 = y12
return sca m13 = y13

return sca d1 = z1
return sca d2 = z2
return sca d3 = z3
return sca d4 = z4
return sca d5 = z5
return sca d6 = z6
return sca d7 = z7
return sca d8 = z8
return sca d9 = z9
return sca d10 = z10
return sca d11 = z11
return sca d12 = z12
return sca d13 = z13

restore
end

bootstrap ///
f1=r(f1) f2=r(f2) f3=r(f3) f4=r(f4) f5=r(f5) f6=r(f6) f7=r(f7)   ///
f8=r(f8) f9=r(f9) f10=r(f10) f11=r(f11) f12=r(f12) f13=r(f13)
///
m1=r(m1) m2=r(m2) m3=r(m3) m4=r(m4) m5=r(m5)
m6=r(m6) m7=r(m7)   ///
m8=r(m8) m9=r(m9) m10=r(m10) m11=r(m11) m12=r(m12)
m13=r(m13)    ///
d1=r(d1) d2=r(d2) d3=r(d3) d4=r(d4) d5=r(d5) d6=r(d6)
d7=r(d7)   ///
d8=r(d8) d9=r(d9) d10=r(d10) d11=r(d11) d12=r(d12)
d13=r(d13)  , ///
reps(1000) seed(19101940): pacman

estat bootstrap, all

translate @Results "$taxi/mrhf boot2.txt", replace
```

PREPRINT

#### Sensitivity analysis

We constructed a model where frailty was dichotomised using a clinically meaningful cut-point of 0.2.  A marked difference in association with healthcare service use due to medication-related harm was observed (Table S7).

Table S7. Odds ratios (ORs) from logistic regression model (dependent variable: healthcare service use due to medication-related harm)

| | OR | 95%CI | | p-value |
|---|---|---|---|---|
| age | 1.01 | 1.00 | 1.03 | 0.137 |
| gender | 0.64 | 0.48 | 0.84 | 0.001 |
| polypharmacy | 1.07 | 1.04 | 1.11 | < 0.001 |
| frailty (binary) | 1.37 | 1.04 | 1.81 | 0.027 |
| model intercept | 0.07 | 0.02 | 0.34 | 0.001 |

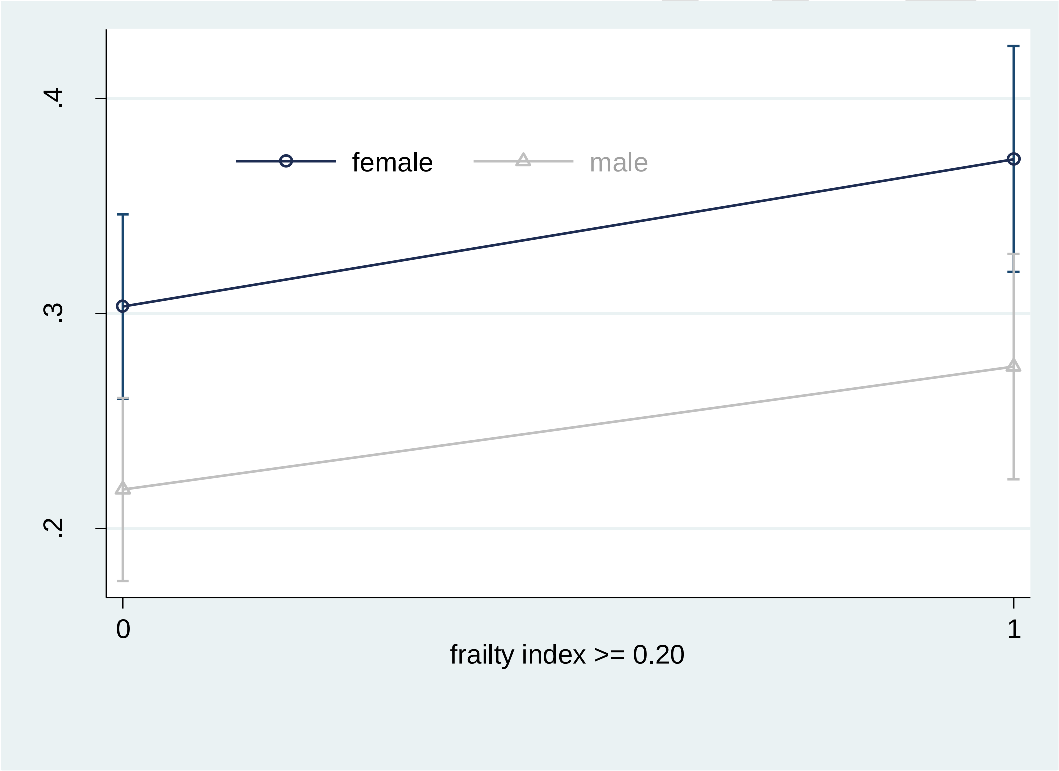

Figure S4 Average marginal estimates (95% confidence intervals) of probabilities of MRH healthcare use when frail or not frail (using 0.2 as the clinically relevant cut-point).
